# Sentinel Event Surveillance to Estimate Total Sars-CoV-2 Infections, United States

**DOI:** 10.1101/2020.03.17.20037648

**Authors:** Andrew A. Lover, Thomas McAndrew

**Affiliations:** Department of Biostatistics and Epidemiology, University of Massachusetts- Amherst, Amherst, MA

**Keywords:** Infectious disease surveillance, Emerging pathogens, Outbreak science, Epidemiology of epidemics

## Abstract

Human infections with a novel coronavirus (SARS-CoV-2) were first identified via syndromic surveillance in December of 2019 in Wuhan China. Since identification, infections (coronavirus disease-2019; COVID-19) caused by this novel pathogen have spread globally, with more than 180,000 confirmed cases as of March 16, 2020. Effective public health interventions, including social distancing, contact tracing, and isolation/quarantine rely on the rapid and accurate identification of confirmed cases. However, testing capacity (having sufficient tests and laboratory throughput) to support these non-pharmaceutical interventions remains a challenge for containment and mitigation of COVID-19 infections.

We undertook a sentinel event strategy (where single health events signal emerging trends) to estimate the incidence of COVID-19 in the US. Data from a recent national conference, the Conservative Political Action Conference, (CPAC) near Washington, DC and from the outbreak in Wuhan, China were used to fit a simple exponential growth model to estimate the total number of incident SARS-CoV-2 infections in the United States on March 1, 2020, and to forecast subsequent infections potentially undetected by current testing strategies. Our analysis and forecasting estimates a total of **54,100** SARS-CoV-2 infections (80 % CI 5,600 to 125,300) have occurred in the United States to March 12, 2020.

Our forecast predicts that a very substantial number of infections are undetected, and without extensive and far-reaching non-pharmaceutical interventions, the number of infections should be expected to grow at an exponential rate.

## 1 Introduction

To date, testing for COVID-19 cases (clinical and otherwise) has been limited in the US. Hospitals and healthcare facilities that do have tests have limited the testing criteria to those with highest potential risk, to optimize the use of scarce resources [1]. Where widespread testing has occurred, increased prevalences in high-risk groups have been found [2, 3].

Evidence-based public health programming depends on data-driven estimates of disease burden to develop interventions. As such, accurate estimate of the underlying burden of SARS-CoV-2 infection within the US can provide critical information for hospital administrators, county- and state-level Departments of Health, and the US Centers for Disease Control and Prevention (CDC) to optimize and prioritize resource allocation.

The Conservative Political Action Conference (CPAC), that took place Feb. 26-29, 2020 outside Washington DC is a large annual event that draws approximately 20,000 participants from across the entire US, thus serving as a representative national sample. A single person was reported as ‘presumptive positive’ for COVID-19 on Mar 7, 2020, and exposure was reported to be prior to the CPAC event [4].

The point incidence of COVID-19 in this defined cohort was used to estimate the national incidence during this period, and subsequent forecasting of current SARS-CoV-2 infections across the entire US. The number of attendees and number of confirmed cases were used to fit a simple exponential growth model to estimate the total COVID-19 infections per day throughout the United States up to March 15th, 2020.

## 2 Overview of Data and Methods

This analysis proceeded in three stages. Firstly, the point incidence of SARS-CoV-2 infection was calculated for the CPAC cohort. Secondly, With an assumption that this group was a near-random sample of Americans, this proportion (daily incidence rate) was then extrapolated to the entire US population (329 million persons) to provide an estimated national daily incidence on March 1, 2020. Finally, this estimate with appropriate error structures, was projected forward in time using epidemiological parameters from the epidemic in Wuhan, PRC to forecast cumulative infections on March 12, 2020.

Data for this analysis were complied from routine sources; press reports from CPAC; and parameters estimated from modeling studies for the early (exponential growth phase) of the epidemic in China. The number of attendees and the number of confirmed COVID-19 cases at CPAC was collected from press reporting. The epidemic doubling time was taken from previous work which estimated from data on the outbreak in Wuhan, China. This doubling time from the Wuhan data was estimated to be 6.4 days, (95 CrI 5.8–7.1), and we used a lognormal distribution for *D* following literature values for other doubling times [5, 6].

## 3 Results and Conclusions

Projecting this total infection burden forward (Fig. 1) using doubling time parameters from Wuhan gives a national cumulative incidence estimate of **54,145** infections (80 % CI 5,647 to 125,274) across the United States up until March 12, 2020. This total includes all infections-asymptomatic, subclinical and clinical cases.

**Figure 1:**
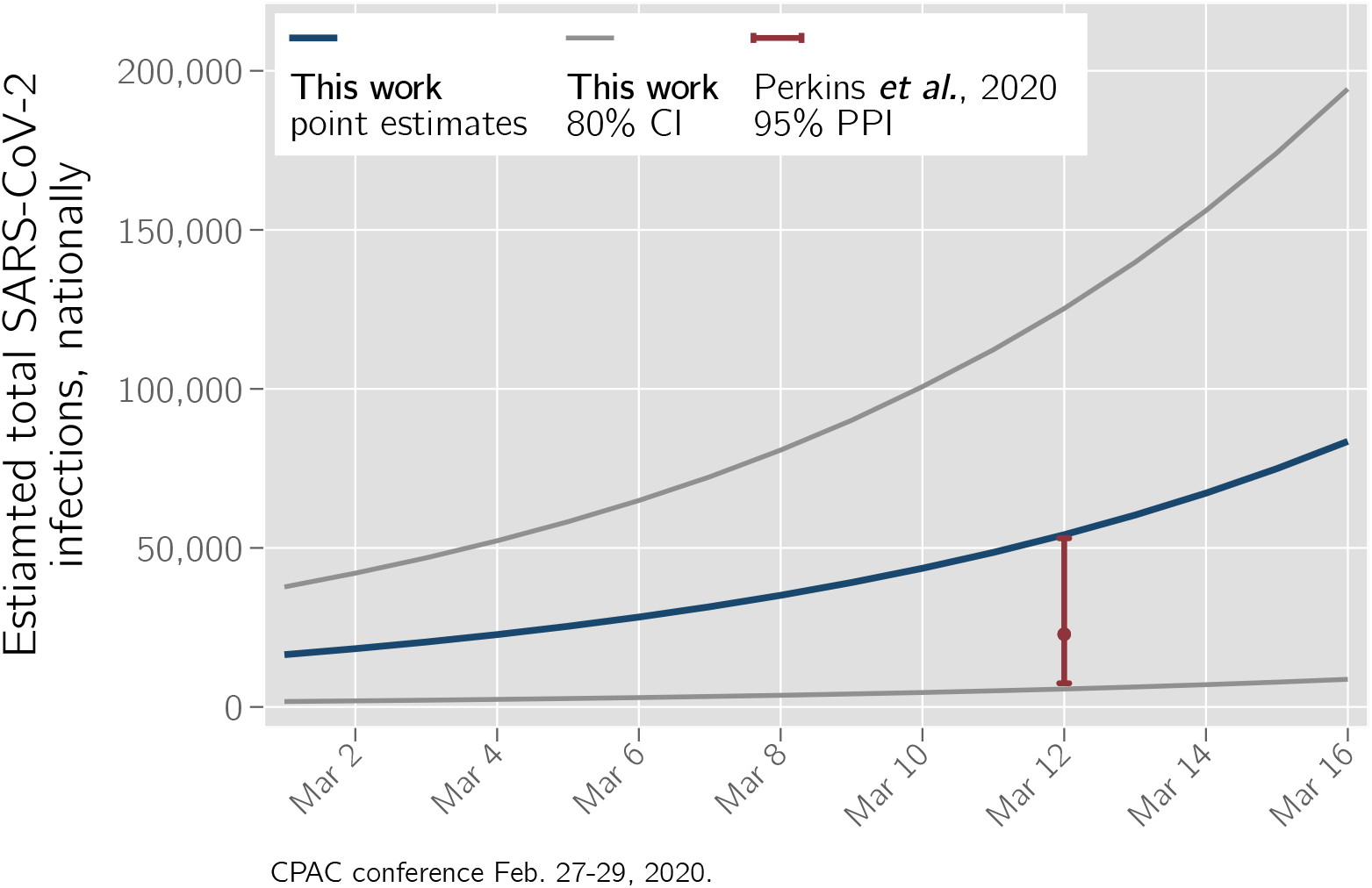
Projected cumulative national SARS-CoV-19 incidence, after March 1, 2020.

This forecasted cumulative number of infections is consistent with those of Perkins *et al*., from independent data sources and with entirely different modeling frameworks, and greatly strengthens the validity of both sets of estimates [7]. The large number of predicted infections compared against the currently reported total confirmed COVID-19 cases in the United States suggest an important proportion of infections may be currently undetected, while entering a phase of potentially very rapid growth of the epidemic.

## 4 Limitations of this analysis

This analysis is not without limitations. Our forecasts are based on parameter estimates using best-available data at the time of analysis to inform public health policy (coined “outbreak science” [8]). Demographic data for CPAC attendees was not available, so we are unable to assess how representative this sentinel event may be for the entire US population. This analysis also relies on simplistic models that assumes a well-mixed population, and uses data derived from transmission scenarios that may not represent transmission within the US. The upper limit of total infections should be interpreted with caution, as small uncertainties early in the projections also accumulate exponentially. Finally, this analysis does not account for spatial heterogeneity and diverse population mixing patterns throughout the United States, which are a critical driver of observed transmission.

## 5 Detailed Methods

### 5.1 The model

We assumed that cumulative infections up to day *t*, denoted by *I*_*t*_, followed an exponential growth model:

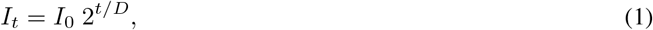

where *I*_0_ is the number of initial infections, *t* is the time, in days, since March 1, 2020, and *D* is the doubling time (in days). Estimates of *I*_0_ and *D* are needed to forecast the cumulative number of infections at time *t*.

### 5.2 Estimating *I*_0_

We estimated total incidence among the US population on March 1st, denoted by *I*_0_, as follows,

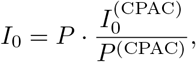

where *P* denotes the size of the US population (329M, [9]), 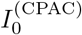 the incidence among CPAC attendees and *P* ^(CPAC)^ the number of CPAC attendees. The parameter *I*_0_ represents the assumed fraction of the US population that is infected with COVID19. The incidence estimated from CPAC was extrapolated to the US population based on the assumption of equal incidence in CPAC attendees and the US population.

The initial relative incidence 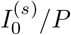 was assumed to follow a beta distribution

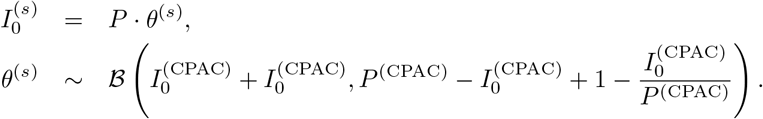

with a prior over *θ*, the probability of infection, equal to a 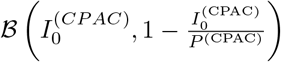 The prior is generated by having observed one sample from the general population with incidence “as observed in the CPAC population”

### 5.3 Estimating *D*

The doubling time of the disease was taken from previous work using data on the outbreak in Wuhan, China, and was estimated to be 6.4 days, (95 CrI 5.8–7.1) and lognormally distributed [5, 6]

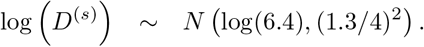

### 5.4 Model-based projections

Point estimates for projected incidence were obtained directly using Eq. 1 and the point estimates of doubling time and initial incidence. Uncertainty in the number of infections was obtained by taking 30, 000 Monte Carlo samples of *I*_0_ and *D* according to their probability densities, and constructing a probability mass function for trajectories, *I*_*t*_,

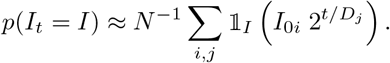

Specifically, the 80% projection interval for *I*_*t*_ is given by the 10^th^ and 90^th^ percentiles of samples 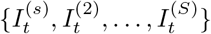. Analysis was performed using R [10] for a 16-day window after March 1, in 1-day intervals. Standard 95% and 80% confidence intervals were constructed across the forecast period.

## Data Availability

All data and statistical analysis code have been uploaded onto GitHub with open access (CC-by).

https://github.com/andrewlover/COVID_19_CPAC_estimation

## 6 Acknowledgements

The authors are grateful for feedback and insightful suggestions from Nick Reich and Leontine Alkema (UMass-Amherst).

## Revision History

| Revision | Date | Author(s) | Description |
| --- | --- | --- | --- |
| 1.0 | 17.1.20 | AAL, TMc | created |
| 1.1 | 17.1.20 | AAL | revised plot legends |

## Notes

### Competing Interest Statement

The authors have declared no competing interest.

### Funding Statement

No specific funding was used for this work.

